# Variants in Vitamin D-related Genes and Prostate Cancer Risk in Black Men

**DOI:** 10.1101/2024.06.29.24309698

**Authors:** Tracy M. Layne, Joseph H. Rothstein, Xiaoyu Song, Shaneda Warren Andersen, Emma K.T. Benn, Weiva Sieh, Robert J. Klein

## Abstract

**BACKGROUND:** The relationship between vitamin D and prostate cancer has primarily been characterized among White men. However, Black men have higher prostate cancer incidence and mortality rates, chronically low circulating vitamin D levels, and ancestry-specific genetic variants in vitamin D-related genes. Here, we examine six critical genes in the vitamin D pathway and prostate cancer risk in Black men.

**METHODS:** We assessed a total of 69 candidate variants in six genes (*GC, CYP27A1, CYP27B1, CYP24A1, VDR*, and *RXRA*) including functional variants previously associated with prostate cancer and circulating 25(OHD) in White men. Associations with prostate cancer risk were examined using genome-wide association study data for approximately 10,000 prostate cancer cases and 10,000 controls among Black men and over 85,000 cases and 91,000 controls among White men. A statistical significance threshold of 0.000724 was used to account for the 69 variants tested.

**RESULTS:** None of the variants examined were significantly associated with prostate cancer risk among Black men after multiple comparison adjustment. Four variants tested P<0.05 in Black men, including two in *RXRA* (rs41400444 OR=1.09, 95% CI: 1.01-1.17, *P* = 0.024 and rs10881574 OR = 0.93, 0.87-1.00, *P* = 0.046) and two in *VDR* (rs2853563 OR = 1.07, 1.01-1.13, *P* = 0.017 and rs1156882 OR = 1.06, 1.00-1.12, *P* = 0.045). Two variants in *VDR* were also positively associated with risk in White men (rs11568820 OR = 1.04, 1.02-1.06, *P* = 0.00024 and rs4516035 OR = 1.03, 1.01-1.04, *P* = 0.00055).

**CONCLUSION:** We observed suggestive non-significant associations between genetic variants in *RXRA* and *VDR* and prostate cancer risk in Black men. Future research exploring the relationship of vitamin D with cancer risk in Black men will need larger sample sizes to identify ancestry-specific variants relevant to risk in this population.

## 1. INTRODUCTION

Research suggests that the circulating form of vitamin D used to define status, 25-hydroxyvitamin D (25(OH)D),^1^ is positively associated with prostate cancer incidence and aggressive disease,^2^ though inversely associated with fatal disease.^3^ These findings are based on extensive large-scale research among European ancestry (“White”) populations.^2, 3^ Conversely, relatively smaller studies among African ancestry (“Black”) populations suggests protective associations between 25(OH)D with prostate cancer incidence^4, 5^ and survival among Black men.^6^ Potential Black-White variation in the vitamin D-prostate cancer association is meaningful as Black men are at high risk for suboptimal vitamin D status^4^ and poor prostate cancer outcomes. ^5^

### 1.1 Black-White Differences in Prostate Cancer Risk and Vitamin D Status

Black men have a 73% higher risk of developing prostate cancer and a more than two-fold higher risk of dying from their disease compared to White men.^5^ In terms of vitamin D status, Black populations are more likely to fall into deficient categories regardless of the definition,^4, 6^ such that non-Hispanic Black participants in a national study were found to have the highest prevalence of severe and moderate vitamin D deficiency compared to other racial/ethnic groups.^4^ Lower circulating 25(OH)D in Black populations is largely driven by higher melanin content in darker skin that slows vitamin D production via exposure to solar ultra-violet B radiation -- the primary source of vitamin D.^1, 7^

### 1.2 Vitamin D Synthesis, Anticancer Activity, and Genomic Impact

The biologically active form of vitamin D, 1,25-dihydroxyvitamin D or 1,25(OH)_2_D_3_, is a pleiotropic hormone with potential physiologic influence on multiple tissues,^8^ and multiple genes and pathways mediate its anti-cancer activity.^9^ Activation of biologic vitamin D is dependent on several steps, including hydroxylation to convert precursor vitamin D into 25(OH)D in the liver,^10^ and to convert 25(OH)D into hormonally active 1,25(OH)_2_D_3_ in the kidney^11^ and at extra-renal sites such as the prostate.^12^ Notably, androgen receptor expression can be regulated by 1,25(OH)_2_D_3_.^13^ 1,25(OH)_2_D_3_ becomes biologically active when it enters the nucleus^8^ and binds to the vitamin D receptor (VDR), a ligand-activated transcription factor^7^ that binds to DNA forming a heterodimer with the retinoid X receptor (RXR).^7, 14^ While various RXR isotypes (i.e., beta and gamma) can form this heterodimer, VDR preferentially associates with RXR alpha (RXRA) to direct transcription.^14^ The 1,25(OH)_2_D-VDR-RXR complex interacts with vitamin D response elements in regulatory regions of target genes, which recruits other transcription factors responsible for calcium and phosphorus homeostasis, but also cell proliferation, apoptosis, differentiation, angiogenesis, invasion and metastasis, and immune response.^7, 15^ VDR, specifically, has been found to occupy over 2,700 genomic positions,^16^ representing 3-5% of the human genome regulatory regions.^7^ VDR is present in normal and malignant prostate tissue^17^ along with the vitamin D-related enzymes needed for vitamin D production, transport, action, and metabolism.^18^ The genomic impact of vitamin D, as facilitated by VDR,^7^ is evident in the 229 genes found to undergo significant changes in expression in response to vitamin D.^16^ Research dedicated to understanding vitamin D’s transcriptional regulation of signaling pathways relevant to prostate cancer in Black men is needed.

### 1.3 Ancestry-specific Genomic Profile and Activity of Vitamin D

Vitamin D metabolites,^12^ carrier protein,^19^ and VDR^20^ have all been found to exhibit ancestry-specific genomic profiles and activity. Both absolute and relative measures of biologically active 1,25(OH)_2_D_3_ were found to be positively associated with West African genetic ancestry.^21^ The vitamin D binding protein (DBP), the primary carrier responsible for transporting circulating 25(OH)D to target tissues, has a well-documented ancestry-specific profile.^19^ DBP is encoded by the growth-specific component gene (*GC*),^19^and binds at least 85% of circulating 25(OH)D.^19^ The combination of two functional polymorphisms in GC, rs4588 and rs7041, result in the three common alleles of the GC protein: Gc1s, Gc1f, and Gc2.^22^ The resultant different versions of the protein, or GC isoforms, differ in their affinity for circulating 25(OH)D; notably, there are well-established Black-White differences in the distribution of these *GC* variants and associated isoforms.^19, 23^ There are also known *VDR* single nucleotide polymorphism (SNP) genotype differences by ancestry,^24^ including a transcriptionally active variant allele more common in African ancestry populations^25^ that may be associated with prostate cancer incidence and mortality in Black men.^20^

The persistent gap in vitamin D-prostate cancer research focused on Black men likely perpetuates existing disparities for a potential risk factor that is *modifiable*. Given the paucity of such research, we examined the role of genes in the vitamin D pathway in relation to prostate cancer among Black men.

## 2. MATERIALS AND METHODS

### 2.1 Vitamin D Pathway Genes and Genetic Variants

We examined genetic variants in the following six critical genes in the vitamin D pathway: 1) *VDR*; 2) *RXRA*; 3) growth-specific component (*GC*), which encodes DBP;^19^ 4) *CYP27A1*, which encodes the enzyme that converts precursor vitamin D to 25(OH)D in the liver; 5) *CYP27B1*, which encodes the enzyme that converts 25(OH)D to hormonally active vitamin 1,25(OH)_2_D_3_ in the kidney; and 6) *CYP24A1*, which encodes the enzyme that degrades 25(OH)D and 1,25(OH)_2_D_3_. ^7,15,26^ From these genes we selected 69 variants (**Appendix Table A.1**) that were: 1) previously studied in relation to prostate cancer incidence and/or mortality, including commonly studied functional variants^27^ such as those known to affect VDR’s affinity for binding biologically active vitamin D^3^, *VDR* variants (Fok1, Apa1, CDX-2, Bsm1, and Taq1) that influence VDR expression in the prostate,^25^ as well as other variants reported to be associated at P<0.05^28, 29^ that were primarily examined in White men^28, 30, 31^ and in a few studies of Black men;^17, 32^ and lastly 2) variants that reached genome-wide significance in genome-wide association studies (GWAS) of circulating 25(OH)D in White populations.^33, 34^Of these variants, only those with summary statistic data available for both ancestral groups were considered.

We used summary statistics data from a previously published large-scale GWAS meta-analysis that included over 10,000 prostate cancer cases and 10,000 controls among Black men and over 85,000 cases and 91,0000 controls among White men.^35^ Data was downloaded from the Database of Genotypes and Phenotype (dbGaP) under accession phs001120.v2.p2. Our primary analysis examined vitamin D genetic variants in relation to prostate cancer risk in Black men, and a secondary analysis compared these findings to those in White men.

### 2.2 Statistical Analysis

A statistical significance threshold of 0.000724 (0.05/69 candidate variants) was used to adjust for multiple comparisons. We used this Bonferroni correction method which is standard, though conservative due to potential correlation among variants within genes. Fisher’s exact test was used to examine allele frequency differences across populations (European vs. African ancestry; and across African subpopulations). To further investigate findings for genetic variants in or near *RXRA* and *VDR*, we used an integrated haplotype score (iHS) to assess whether regions around variants of interest show evidence of recent selection within African or European populations. iHS uses a standardized measure of the extent of homozygosity around a site of interest and compares haplotypes containing the derived allele to those containing the ancestral allele in order to assess the evidence for recent selection.^36^ We used the Human Genome Diversity Project iHS track of the UCSC Genome Browser (build hg18)^37^to retrieve iHS scores computed by Pickrell et al.^38^ in Bantu-speaking and European populations. Within African populations, Bantu-specifics scores were computed in order to represent a reasonably homogeneous African population for the detection of recent selection. iHS scores are provided in the form of –log10 of an empirical P-value, which is computed relative to the rest of the genome.^39^ Additionally, gene variant findings were examined based on visual annotation and visualization of linkage disequilibrium using LocusZoom.^40^

## 3. RESULTS

None of the examined 69 variants from the six vitamin D pathway genes were significantly associated with prostate cancer risk among Black men after adjustment for multiple comparisons (**Table A.1**). Four variants were found to be associated at P <0.05 in Black men **Table 1**. These variants include two in *RXRA* [rs41400444 OR=1.09, p =0.024 and rs10881574 OR = 0.93, p =0.04556] and two in *VDR* [rs2853563 OR = 1.07, p= 0.01726 and rs11568820 OR = 1.06, p = 0.0495]. For comparison, among these four nominally significant variants in Black men, rs11568820 was the only variant significantly associated with risk among White men after correction for multiple comparisons (rs11568820 OR = 1.04, p = 0.00024) despite their considerably larger sample size relative to Black men in the study.

**TABLE 1.** Four Vitamin D Pathway Genetic Variants Associated with Prostate Cancer Risk at P<0.05 in Black Men (10,368 cases and 10,986 controls) with comparison to results in White Men (85,554 cases and 91,972 controls).

| Chr | SNP | GRCh37<br>Position | EA | Race | EAF | OR | LCL | UCL | P-value |
| --- | --- | --- | --- | --- | --- | --- | --- | --- | --- |
| RXRA |  |  |  |  |  |  |  |  |  |
| 9 | rs41400444 | 137231651 | T | Black | 0.10 | 1.09 | 1.01 | 1.17 | 0.0242 |
|  |  |  |  | White | 0.09 | 1.01 | 0.98 | 1.04 | 0.7105 |
|  | rs10881574 | 137205222 | T | Black | 0.90 | 0.93 | 0.87 | 1.00 | 0.0456 |
|  |  |  |  | White | 0.92 | 0.99 | 0.96 | 1.02 | 0.5187 |
| VDR |  |  |  |  |  |  |  |  |  |
| 12 | rs2853563 | 48235738 | T | Black | 0.16 | 1.07 | 1.01 | 1.13 | 0.0173 |
|  |  |  |  | White | 0.04 | 1.01 | 0.97 | 1.05 | 0.5028 |
|  | rs11568820 --<br>Cdx2 | 48302545 | T | Black | 0.79 | 1.06 | 1.00 | 1.12 | 0.0495 |
|  |  |  |  | White | 0.20 | 1.04 | 1.02 | 1.05 | 0.0002 |
| Chr = chromosome; SNP = single nucleotide polymorphism; GRCh37 = Genome Reference Consortium Human Build 37; EA = effect allele (*among controls*); EAF = effect allele frequency; VDR = vitamin D receptor; RXRA = retinoid X receptor alpha |  |  |  |  |  |  |  |  |  |

Among these four variants, the effect allele frequency for rs11568820 differed substantially between African (0.79) and European (0.20) ancestry populations based on the Genome Aggregation Database (**Table 1**).^41^ Furthermore, a global test for differences among five African subpopulations revealed significant differences in allele frequencies for rs11568820 (Fisher’s exact P-value = 0.000691). In our assessment of recent selection based on iHS score, *VDR* rs2853563 had a score of 0 (p=1.0) in both Bantus and Europeans, and rs11568820 had scores of 0.486809 (p=0.33) and 0 (p=1.0) in Bantus and Europeans, respectively, arguing against recent selection at these variants. Similarly, the variants in (rs41400444) or near (rs10881574) *RXRA* had iHS scores of 0 (p=1.0) in Bantu populations and 0.737046 (p=0.18) in European populations. Thus, we do not observe any evidence supporting recent selection around these variants in Bantu or European populations.

To place these variants in their genomic context, we examined the ancestry-specific linkage disequilibrium (LD) pattern within 200kb around each variant. The visualization for VDR variant rs11568820 is depicted in **Figure 1** given that it demonstrated nominally significant results in Black men and significant results in White men. We note that LD patterns generally show weaker LD with rs11568820 in African (**Figure 1A**) vs. European ancestry (**Figure 1B**) populations. In both African and European ancestry populations, other SNPs in this region were more highly significantly associated with prostate cancer risk than rs11568820 suggesting that it is unlikely to be the causal variant. For the remaining three variants, different ancestry-specific LD patterns were also present (**Appendix Figure A.1**), along with more highly significant SNPs in the region, suggesting that none of the nominally significant variants are likely to be causal.

**Figure 1:**
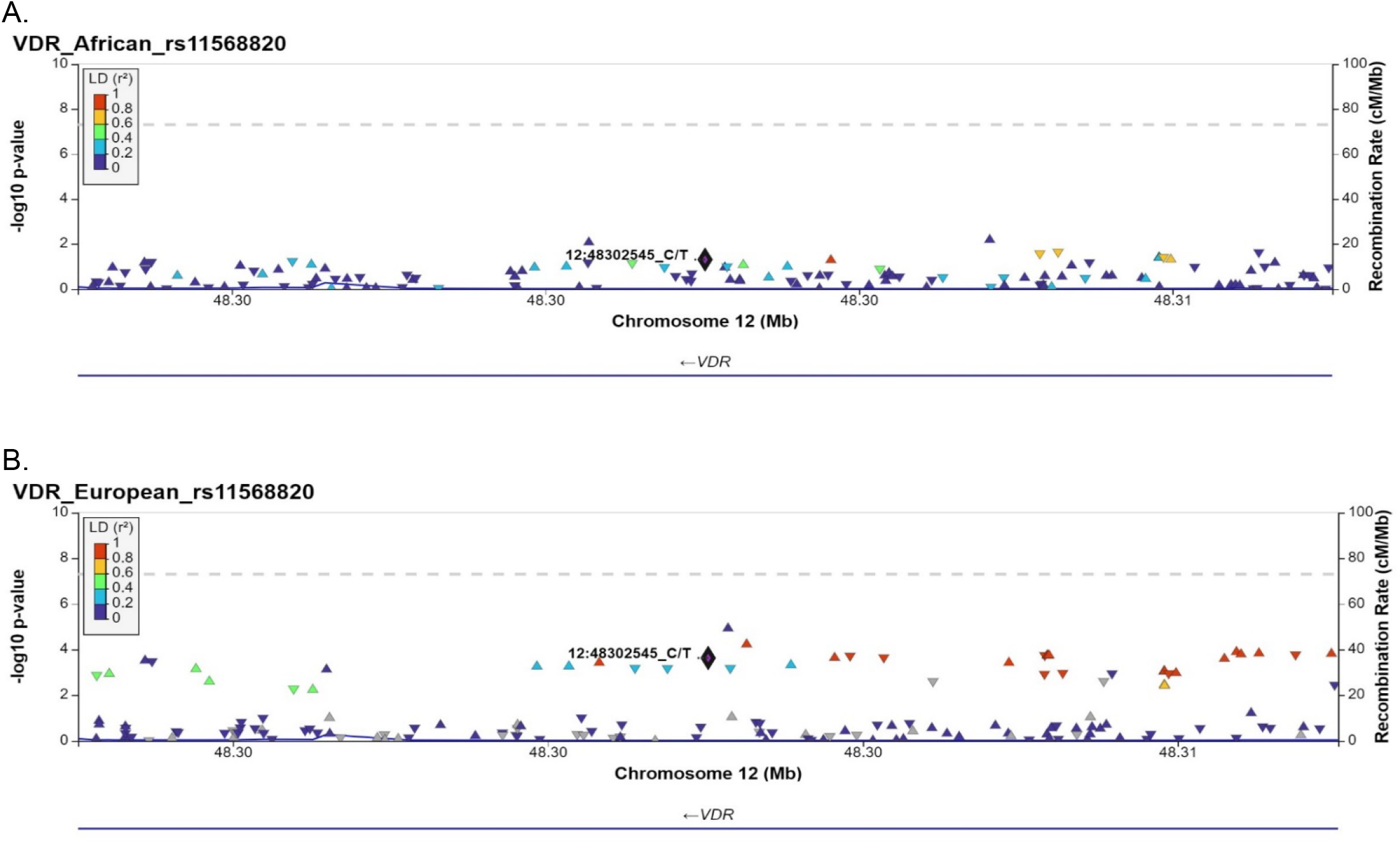
Locus Zoom plot of VDR variant rs11568820 (± 200kb) on chromosome 12 [GRCh37 position: 48302545] for African (A) and European (B) ancestry populations. The Y-axis represents the −log_10_(p value) for SNPs in the region (triangles) and the color of each triangle represents the degree of linkage disequilibrium (LD) with rs11568820.

## 4. DISCUSSION

In our analysis of genetic variants in six candidate genes in the vitamin D pathway, we observed suggestive associations between variants in the genes *RXRA* and *VDR* with the risk of prostate cancer among Black men. We also observed LD patterns that both appear to differ between African and European ancestry populations, and that suggest these are likely not causal variants, particularly for the commonly studied functional *VDR* variant rs11568820. However, these specific genes are central to vitamin D’s biologic activity and have been linked to prostate cancer. Namely, VDR may mediate the role of testosterone and vitamin D in prostate cancer,^25^ and operate differently in prostate cancer among Black and White men;^24, 42^ differences that may arise, in part, due to ancestry-specific differences in genetic variant allele frequencies.^20, 24^Additionally, suppression of *RXRA*, a nuclear receptor that mediates retinoic acid-mediated gene activation, was identified as a novel target by microRNA-191 to promote radiation resistant prostate cancer.^43^

### 4.1 Prior Findings for the Four Highlighted Variants in *RXRA and VDR*

In the current study, all four of the highlighted variants demonstrated suggestive positive associations with prostate cancer risk in Black men. Data from a large cohort consortium study of genetic risk factors and risk of breast and prostate cancer, observed null associations for the two variants on *RXRA* (rs41400444 per minor allele OR=1.05, 95%CI:0.72-1.52; rs10881574 OR=0.98, 95%CI:0.74-1.31) and fatal prostate cancer in White men based on 496 fatal cases and 2,986 matched controls without a previous history of prostate cancer.^44^ Our findings also include the *VDR* variant rs11568820, which is known as CDX-2 due to its influence on the affinity of the CDX-2 transcription factor for the VDR promoter region.^45^ Additionally, calcium absorption in the small intestine is regulated by VDR whose expression is influenced by tissue-specific transcription factors, such as CDX-2.^25, 46^ The *VDR* promoter region contains the CDX-2 A/G polymorphism which binds more efficiently to A than the G allele, and as a consequence the A allele is associated with enhanced transcriptional activity,^25^ including for genes regulating calcium absorption which can influence carcinogenesis in prostate cells.^45^ The high activity A allele is more common in African ancestry populations and was hypothesized to influence incidence and mortality in Black men from a population-based case-control study.^20^ A 2016 meta-analysis of *VDR* polymorphisms including CDX-2, found no association with prostate cancer risk overall, although borderline associations were found in the single study of Black men (Homozygote AA vs. GG OR =1.80, 0.97-3.32, p=0.06).^25^ Rowland et al. found that low calcium intake was associated with reduced prostate cancer risk in Black and White men with the CDX-2 low intestinal calcium absorption genotype.^47^ While a different study in Black men observed that risk of advanced prostate cancer decreased with increasing number of the minor CDX-2 G allele, and the relationship was modified by calcium intake (p for interaction=0.03) such that the reduction (50% decrease) was only evident in Black men with calcium intake below the median.^20^ In a study of 155 Black men with prostate cancer, including 46 aggressive cases, controlling for CDX-2 in analytical models increased the magnitude of the association between vitamin D deficiency and aggressive disease (OR=3.64, P= 0.05).^45^ In terms of prior research on *VDR* rs2853563, no association was found with prostate-specific antigen concentration, prostate cancer risk, or Gleason score in a study of Black men.^32^

### 4.2 Other *VDR* Variant Prostate Cancer Associations

In the consortium study by Shui et al., several variants in genes in the vitamin D metabolism and signaling pathway examined in the current study (e.g., GC, CYP27A1, and CYP24A1), along with other variants in VDR, were nominally associated with fatal prostate cancer risk in White men.^44^ A novel *VDR* variant -- referred to as “SNP 4” (c.907+75C>T) – identified in a study of Black men (90 cases/91 controls) was associated with reduced risk (OR =0.08, p<0.001) and low Gleason score (β= −0.64, p<0.05). This study also observed that a previously reported SNP (*Apa1* rs7975232) was associated with risk (OR =3.6, p=0.002) and high Gleason score (β=0.32, p<0.05.^17^ None of these variants were associated with risk in the current study.

### 4.3 Ancestry-specific Genomic Activity of Vitamin D

Potential Black-White variation in the VDR-prostate cancer relationship is plausible given prior research noting that race/ethnicity is an effect modifier of *VDR* polymorphisms cancer associations,^24^ and the known *VDR* SNP genotype differences by ancestry.^24^ Qualitatively and quantitatively distinct VDR signaling patterns have been observed in prostate cancer cell lines derived from Black and White men,^42^ with VDR signaling noted to impact prostate cancer and that African genomic ancestry has a distinct influence on VDR genomic binding in this context.^42^ Ancestry-specific differences in genomic activity and cancer associations may also be true of other vitamin D pathway genes such as *GC*. Interestingly, circulating 25(OH)D was found to be protective for overall and lung cancer survival among those with the Gc2 isoform;^48^ the isoform *least* common in Black populations.^19^ That there is African ancestry-specific genomic activity of vitamin D-related genes that influence risk for cancers with disproportionate disease burden in Black populations, and the potential for genomic activity to be altered in a beneficial way with supplementation in a chronically vitamin D deficient group,^18^ is striking.

This speaks to the importance of identifying and accounting for ancestry-specific genetic profiles.^49, 50^ The absence of such detail is said to underlie the lack of replication of genetic association studies due to differences in LD patterns across ancestral groups which are inadequately capturing causal variants by tagging variants identified from a single population.^50^ A lack of replication has also been linked to differences in genetic architecture related to ancestry-specific variation and allele frequency differences stemming from genetic drift and/or selection.^50^ Related to this, ancestry-concordant models (e.g., populations with similar allele frequencies and accurate assessment of linkage disequilibrium patterns) are noted to be useful for improving the power for gene discovery in the context of transcriptome wide association studies aimed at identifying traits associated genes.^49^

### 4.4 Challenges Studying Genomic Activity of Vitamin D and Prostate Cancer in Black Men

While our findings are based on large-scale data from across multiple studies, specific examinations of genetic variants in vitamin D pathway genes and prostate cancer risk in Black men may have been hampered by common challenges with such group-specific analyses. Specifically, despite having approximately, 10,000 cases and 10,000 controls, our analysis may have been underpowered to detect true associations, a common cause of false-negative or weak associations in studies of Black populations.^3, 51^ Additionally, an inaccurate accounting of the true population genetic structure may have also limited our analysis,^49, 50^ along with admixture in Black populations which can lead to inaccuracies (false-positive or false-negative), and residual confounding even when global ancestry is accounted for.^52^

## 5. CONCLUSION

We observed suggestive non-significant associations between genetic variants in critical vitamin D-related genes – the genes encoding the vitamin D receptor and the retinoid X receptor which are critical to the genomic activity underlying vitamin D’s impact on health, including prostate cancer. Given the investigated genetic variants on these genes were identified largely based on research in White men, future research is needed with large-scale individual-level data from Black populations to better identify vitamin D genetic variants reflective of the prostate cancer risk profile in Black men. As part of this future work, investment in substantially larger studies among men of African ancestry will help not only overcome the challenges related to small sample size, but also provide opportunities to identify variants relevant to prostate cancer in this high-risk, understudied group.

## Data Availability

All data produced in the present study are available upon reasonable request to the authors.

