## Appendix Figure 1 for "Variants in Vitamin D-related Genes and Prostate Cancer Risk in Black Men"

Figure A.1.1

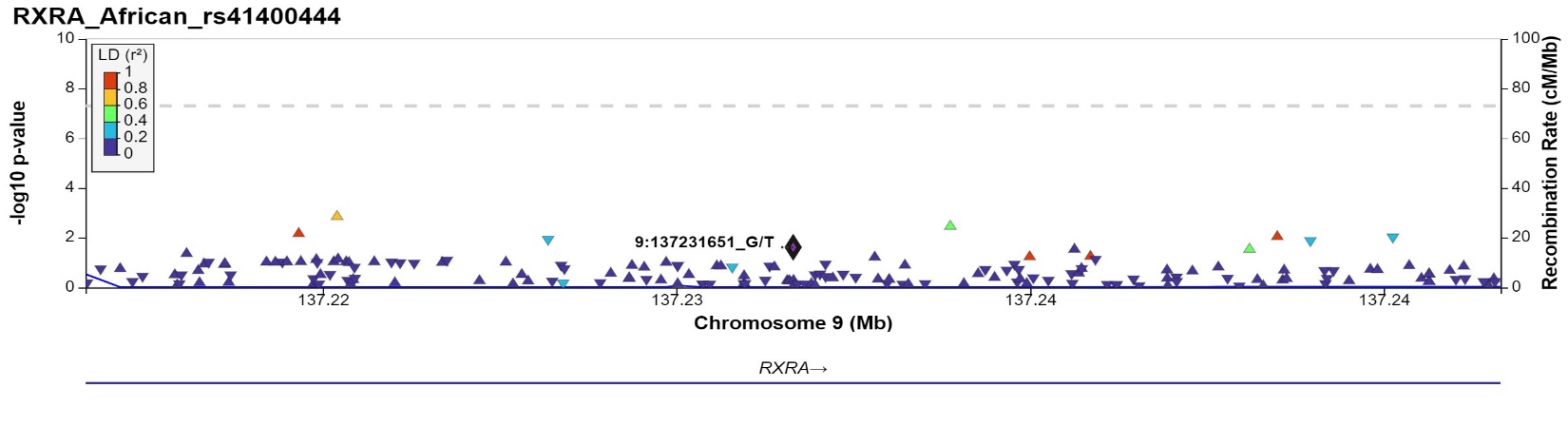

Figure A.1.2

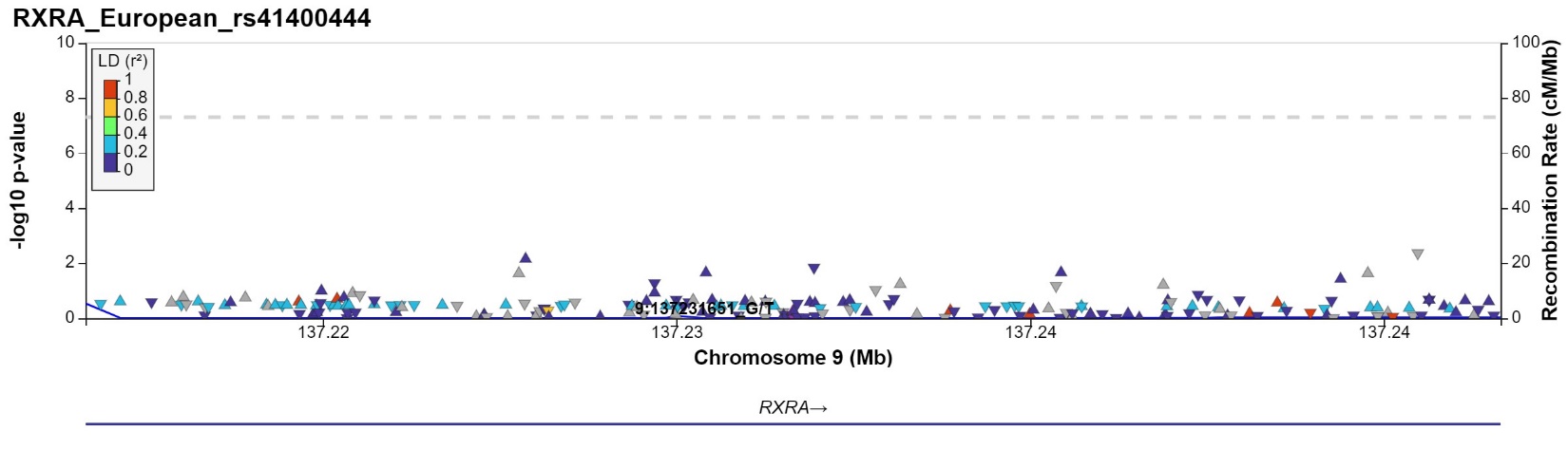

Figure A.1.3

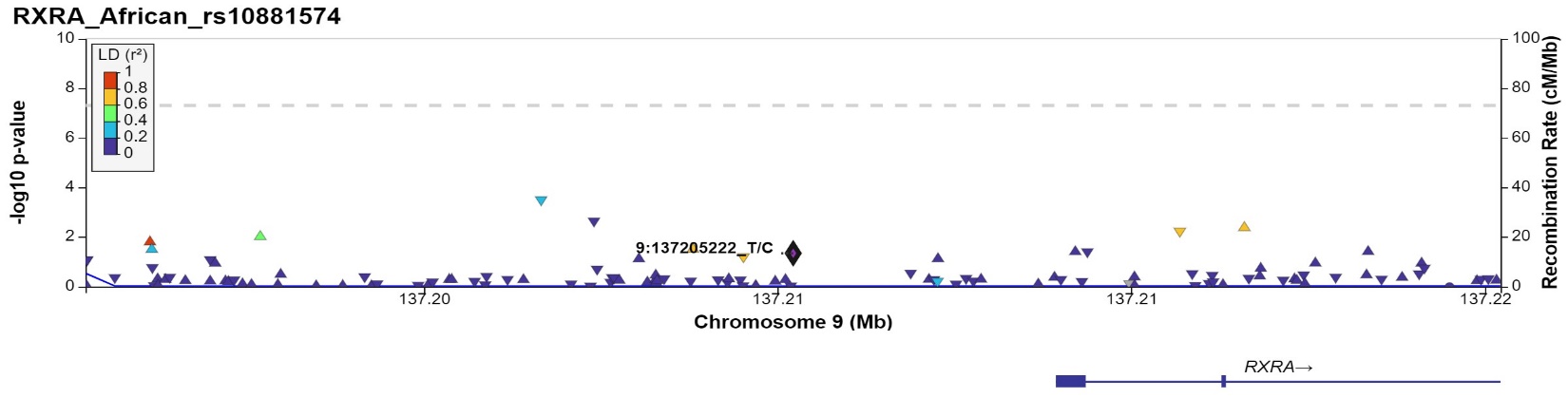

Figure A.1.4

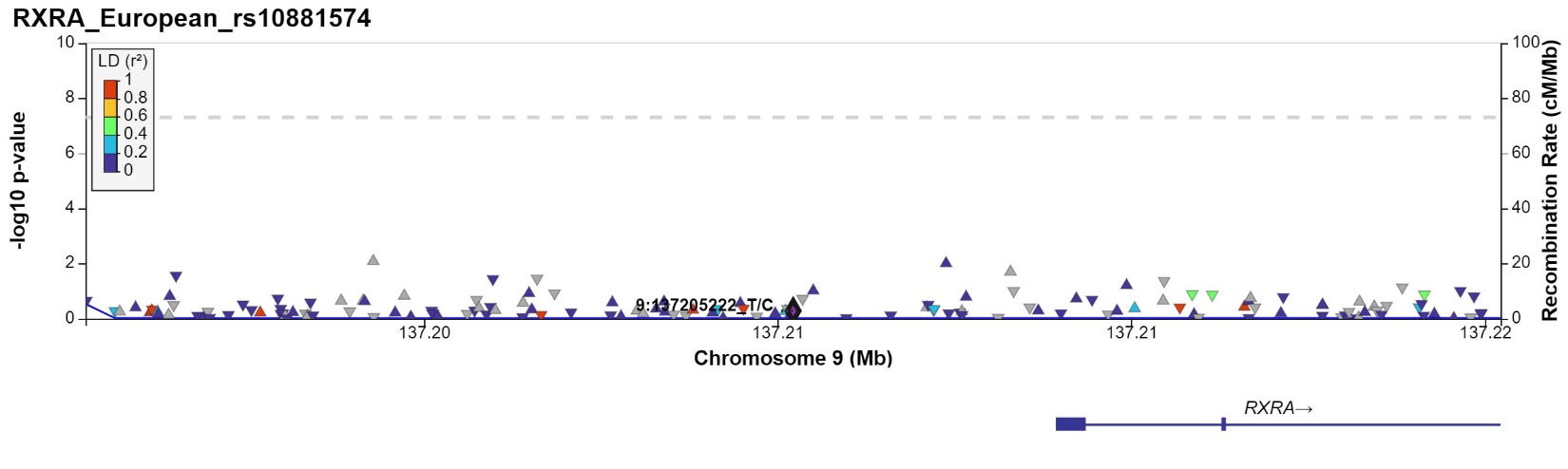

Figure A.1.5

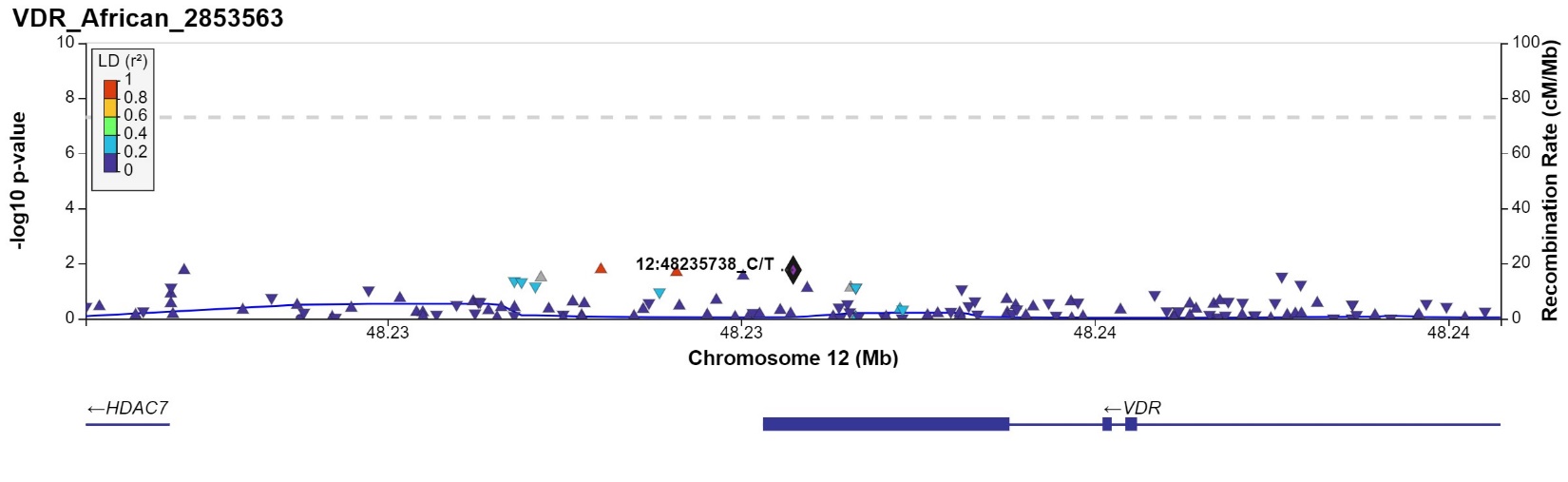

Figure A.1.6

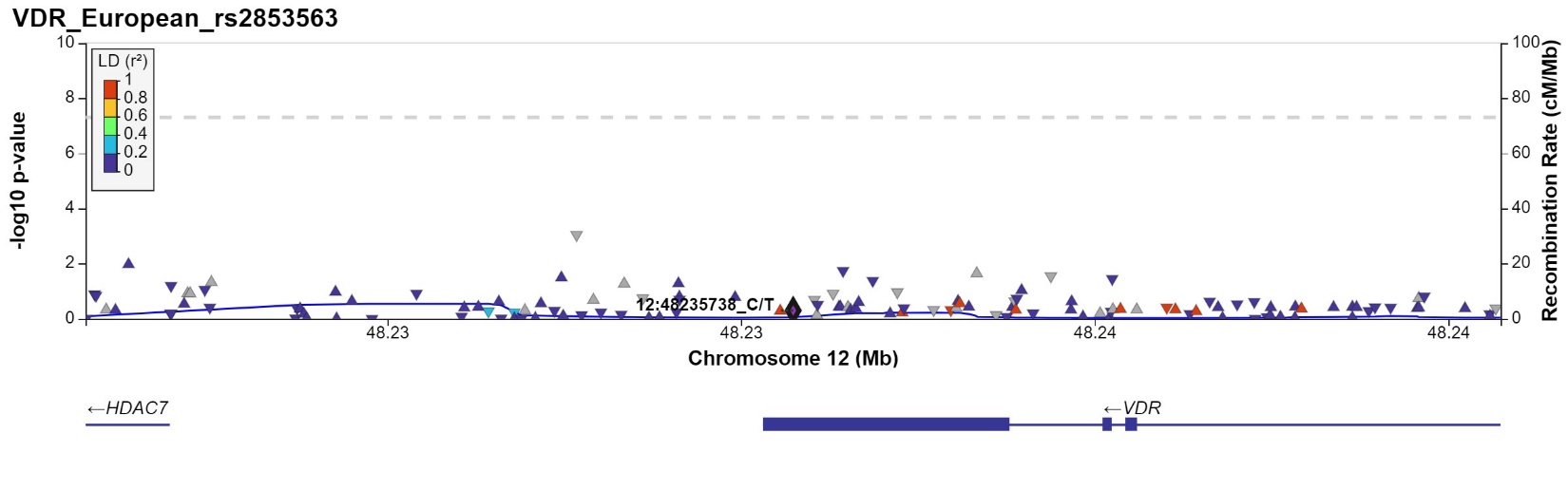

Figure A.1.7

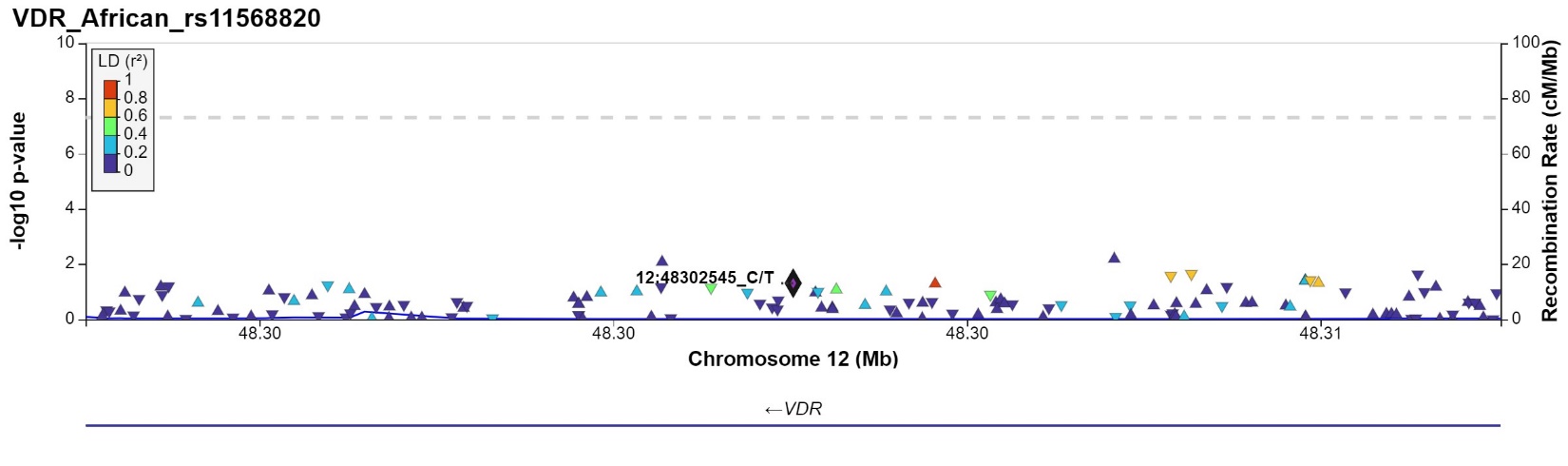

Figure A.1.8

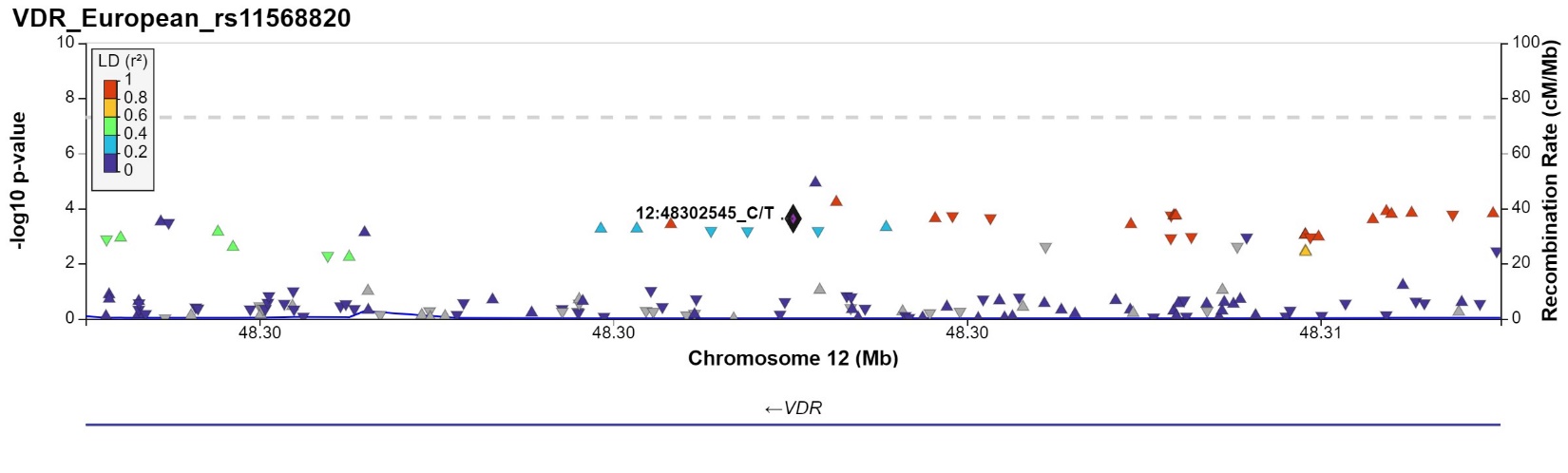

**Figure A.1**: Locus Zoom plot For African and European populations within ± 200kb of the following variants: a) *RXRA* variants rs41400444 on chromosome 9 [GRCh37 position: 137231651], and rs10881574 [GRCh37 position: 137205222]; and VDR variants rs2853563 on chromosome 12 [GRCh37 position: 48235738], and rs11568820 [GRCh37 position: 48302545]. The Y-axis represents the -log_10_(p value) for SNPs in the region (triangles) and the color of each triangle represents the degree of linkage disequilibrium (LD) with rs11568820.
